# Efficacy of electroacupuncture versus transcutaneous electrical acupoints stimulation on subclinical premature ovarian insufficiency: study protocol for a randomized controlled trial

**DOI:** 10.1101/2022.09.19.22280099

**Authors:** Jialing Wang, Linglin Dai, Yani Xu, Xufen Zhang, Yutong Zhang, Ding Tang, Mingjie Zhan, Chao Wang, Zhanglian Wang, Lifang Chen

## Abstract

**Introduction:** “Subclinical stage” of premature ovarian insufficiency (POI) refers to menstrual disorders with FSH levels in the range of 15-25 U/L in women under 40 years old caused by diminished ovarian function. Early intervention of subclinical POI may be able to protect ovarian function more effectively and prevent further development to POI. Currently, no standard-of-care exists for subclinical POI. Previous studies have shown effectiveness of electroacupuncture (EA) on POI and transcutaneous electrical acupoint stimulation (TEAS) on diminished ovarian reserve, but no published studies focus on the treatment of subclinical POI. This study aims to assess whether EA or TEAS will be effective in improving ovarian function at the subclinical stage of POI and preventing the progression.

**Methods:** 114 subclinical POI patients between 25 and 40 years will be randomly assigned to three groups (an EA group, a TEAS group and a waiting for treatment group) in this randomized, controlled, assessor-blinded trial. The treatment will last for three months and the follow-up will last for twelve months. The primary outcomes will be anti-Mullerian hormone (AMH) and follicle-stimulating hormone (FSH). Secondary outcomes will include serum estradiol (E_2_), luteinizing hormone (LH), FSH/LH ratio, the menstrual status assessment, and modified Kupperman Menopausal Index. We will also investigate the incidence of adverse events.

**Trial registration:** Trial was registered in the Chinese Clinical Trail Registry (ChiCTR-2100045598); Pre-results.

## INTRODUCTION

In recent years, with the change in China’s fertility policy, more and more attention has been paid to women’s fertility. Premature ovarian insufficiency (POI), which is considered to be one of the main causes of infertility, is a disease of declining ovarian function that occurs before the age of 40[1]. The incidence of POI is about 1∼3.8% worldwide and shows evidence of younger trends[2-3]. Genetic, immune and environmental factors are common causes of POI. However, more than half of POI is called idiopathic POI with unclear etiology. The range of systemic symptoms resulting from low estrogen causes great physical and mental suffering to patients, including cardiovascular disease, osteoporosis, urogenital atrophy and neurodegenerative disorders[4]. Apart from these diseases, infertility in POI patients is difficult to treat[5-6], making POI one of the most devastating diseases of women in reproductive age. If left untreated, POI can progress to complete ovarian failure within a few years. Based on the value of prevention, the Chinese Expert Consensus on Clinical Treatment of POI specifically defined “subclinical stage” of POI in 2017[7]. A patient with “subclinical stage” of POI has just started to show menstrual disorders, mainly manifesting as sparse or frequent menstruation, with FSH levels in the range of 15-25 U/L. Statistically, 5-10% of POI patients can ovulate and conceive spontaneously[8], which demonstrates that some patients still have the ability to spontaneously recover ovarian function. For this reason, treatment of patients in subclinical POI stage may aid in restoring ovary function. However, hormone replacement therapy (HRT), the first clinical line for POI, has unavoidable contraindications and non-negligible side effects[9-12]. These side effects made HRT sub-optional for the treatment of subclinical POI. Other Other therapeutic modalities such as phytohormones and follicular in vitro activation have not been widely used clinically and their adverse effects have yet to be confirmed by clinical evidence. At the same time, specific targeted therapy for the etiology cannot be performed since the etiology of most patients with POI is unknown.

As a part of traditional Chinese medicine, acupuncture has been used for the treatment of gynecological diseases since thousands of years[13]. Compared to traditional acupuncture, electroacupuncture (EA) and transcutaneous electrical acupoint stimulation (TEAS) use current instead of manual stimulation. They are widely used by clinicians and researchers because of their time-saving and quantifiable advantages. Specifically with the treatment of POI, researchers have found that EA can not only improve menstrual symptoms, ovarian reserve, perimenopausal symptoms, but also demonstrates effects on relieving the negative emotions of patients[14-15]. TEAS was also found effective in treating gynecological disorders, especially infertility, in increasing evidence[16-17]. Several previously published studies have indicated that acupuncture can increase Anti-Müllerian hormone (AMH) and decrease serum follicle-stimulating hormone (FSH) in POI patients[14,18-19]. AMH and FSH can both regulate oocyte competence during aging. Basal AMH and FSH circulatory concentrations have been broadly utilized as in vitro fertilization (IVF) success predictors[20]. The mechanism of acupuncuture treatment on POI may be achieved through regulation of the hypothalamic-pituitary-ovarian axis (HPOA), improve hemodynamics of sexual organ and adjust sex hormone levels[21-23]. However, no previous research has yet focused on the treatment of acupuncture for subclinical POI patients. Therefore, we designed this trial to investigate the efficacy and safety of EA and TEAS options for treating subclinical POI.

## MATERIALS AND METHODS

### Study design

This study will be a single-center, randomized controlled and assessor-blinded trial which will be implemented in The Third Affiliated Hospital of Zhejiang Chinese Medical University. The duration of the whole study will be seventeen months: two months before randomization as baseline, three months of treatment, and twelve months of follow-up. The trial flow chart is presented in **figure 1** and the trial schedule is shown in **table 1**. Any change of the protocol has to be agreed by the entire research team and documented accordingly. In version 2.0 of the trial protocol, the experimental group was treated with EA and the control group was treated with TEAS. However, in the trial process, we found that TEAS also had certain clinical efficacy and was welcomed by the subjects. Since the latest discussion within the research group suggested that TEAS may also be a good treatment for POI, which is worth studying, we added a waiting treatment group to evaluate the therapeutic effect of EA and TEAS on subclinical POI respectively. Also, because of the addition of a waiting treatment group, the recruitment time and the duration of the whole study were changed to better serve the clinical trial. This study protocol was developed according to the Standard Protocol Items: Recommendations for Interventional Trials (SPIRIT) guidelines[24] **(Supplementary file 1)** strictly.

**Figure 1:**
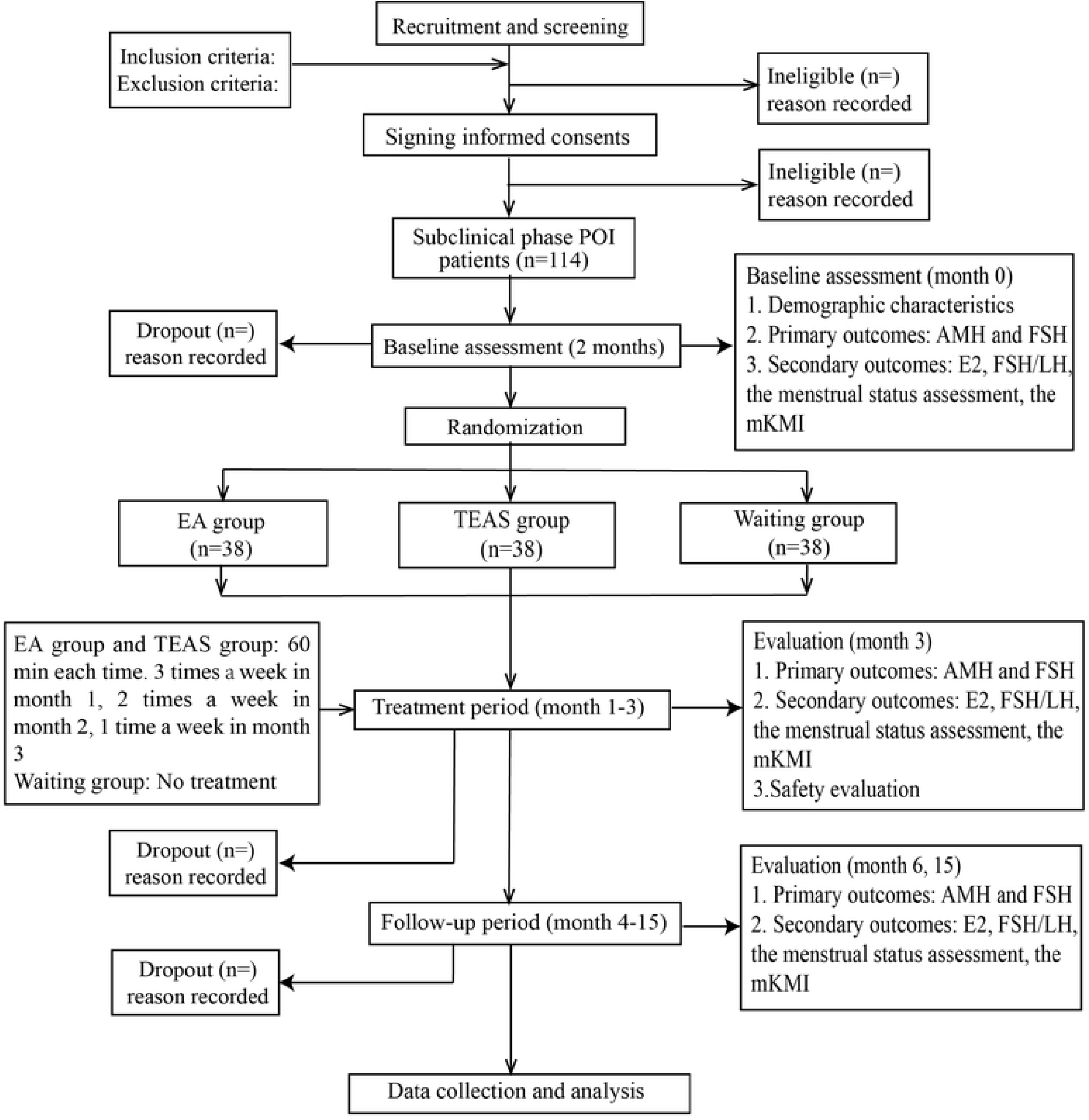

**Table 1.**
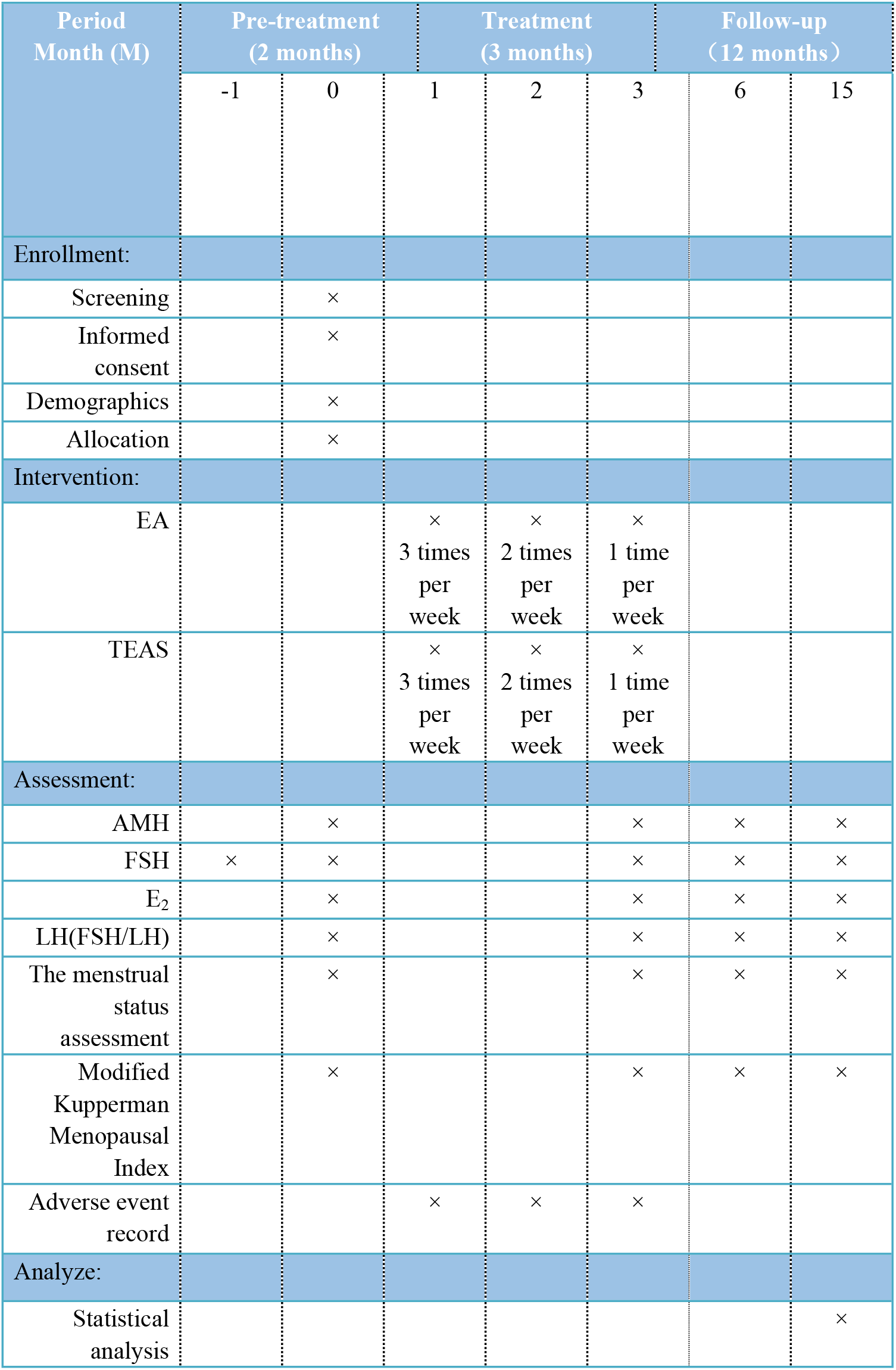
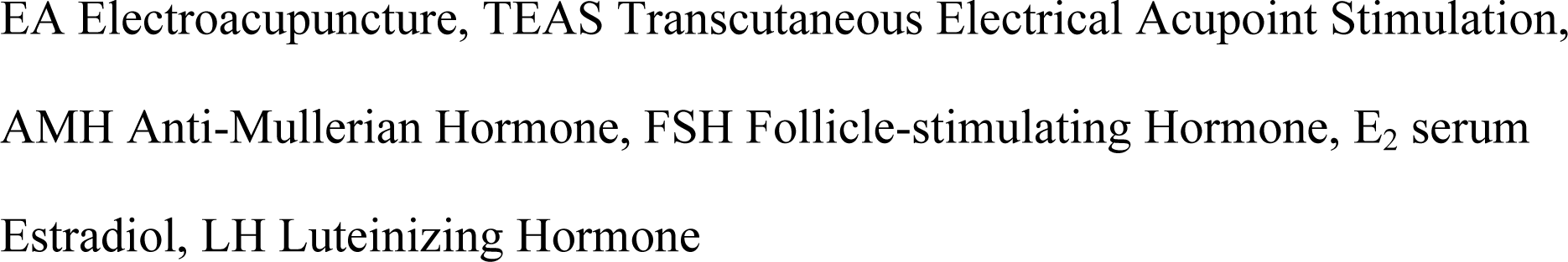
Time schedule

| Period<br>Month (M) | Pre-treatment<br>(2 months) |  | Treatment<br>(3 months) |  |  | Follow-up<br>(12 months) |  |
| --- | --- | --- | --- | --- | --- | --- | --- |
|  | -1 | 0 | 1 | 2 | 3 | 6 | 15 |
| Enrollment: |  |  |  |  |  |  |  |
| Screening |  | × |  |  |  |  |  |
| Informed consent |  | × |  |  |  |  |  |
| Demographics |  | × |  |  |  |  |  |
| Allocation |  | × |  |  |  |  |  |
| Intervention: |  |  |  |  |  |  |  |
| EA |  |  | × | × | × |  |  |
|  |  |  | 3 times<br>per<br>week | 2 times<br>per<br>week | 1 time<br>per<br>week |  |  |
| TEAS |  |  | × | × | × |  |  |
|  |  |  | 3 times<br>per<br>week | 2 times<br>per<br>week | 1 time<br>per<br>week |  |  |
| Assessment: |  |  |  |  |  |  |  |
| AMH |  | × |  |  | × | × | × |
| FSH | × | × |  |  | × | × | × |
| E<sub>2</sub> |  | × |  |  | × | × | × |
| LH(FSH/LH) |  | × |  |  | × | × | × |
| The menstrual status assessment |  | × |  |  | × | × | × |
| Modified Kupperman Menopausal Index |  | × |  |  | × | × | × |
| Adverse event record |  |  | × | × | × |  |  |
| Analyze: |  |  |  |  |  |  |  |
| Statistical analysis |  |  |  |  |  |  | × |

EA Electroacupuncture, TEAS Transcutaneous Electrical Acupoint Stimulation, AMH Anti-Mullerian Hormone, FSH Follicle-stimulating Hormone, E<sub>2</sub> serum Estradiol, LH Luteinizing Hormone

### Ethics and registration

Permission was obtained from the Ethic Committee of The Third Affiliated Hospital of Zhejiang Chinese Medical University (Approved No. ZSLL-KY-2021-004-01) **(Supplementary file 2)** and the trial has been registered in the Chinese Clinical Trail Registry with identifier ChiCTR-2100045598. We will conduct the trail in accordance with the Declaration of Helsinki. Prior to the start of the trial, each subject will be required to sign an informed consent form. During the trial, the subjects’ personal information and data will be stored in an encrypted computer.

### Recruitment and screening

We will place recruitment announcements on the hospital’s WeChat public account, in the hospital lobby and some nearby communities. In the meanwhile, we will ask our gynecologists to help recruit potential subclinical POI patients. After initial confirmation of intent to participate, the coordinator will interview the potential subject face to face, clearly communicate with the subject of the aim, duration and other details of the study, and ask the subject to undergo two FSH tests at our institution. If potential subjects fully meet the inclusion criteria, we will ask him/her to sign a paper informed consent form. The inclusion and exclusion criteria are shown in the following **Table 2**. Recruitment will be open from March 20^th^, 2021 to December 1^st^, 2022.

**Table 2.**
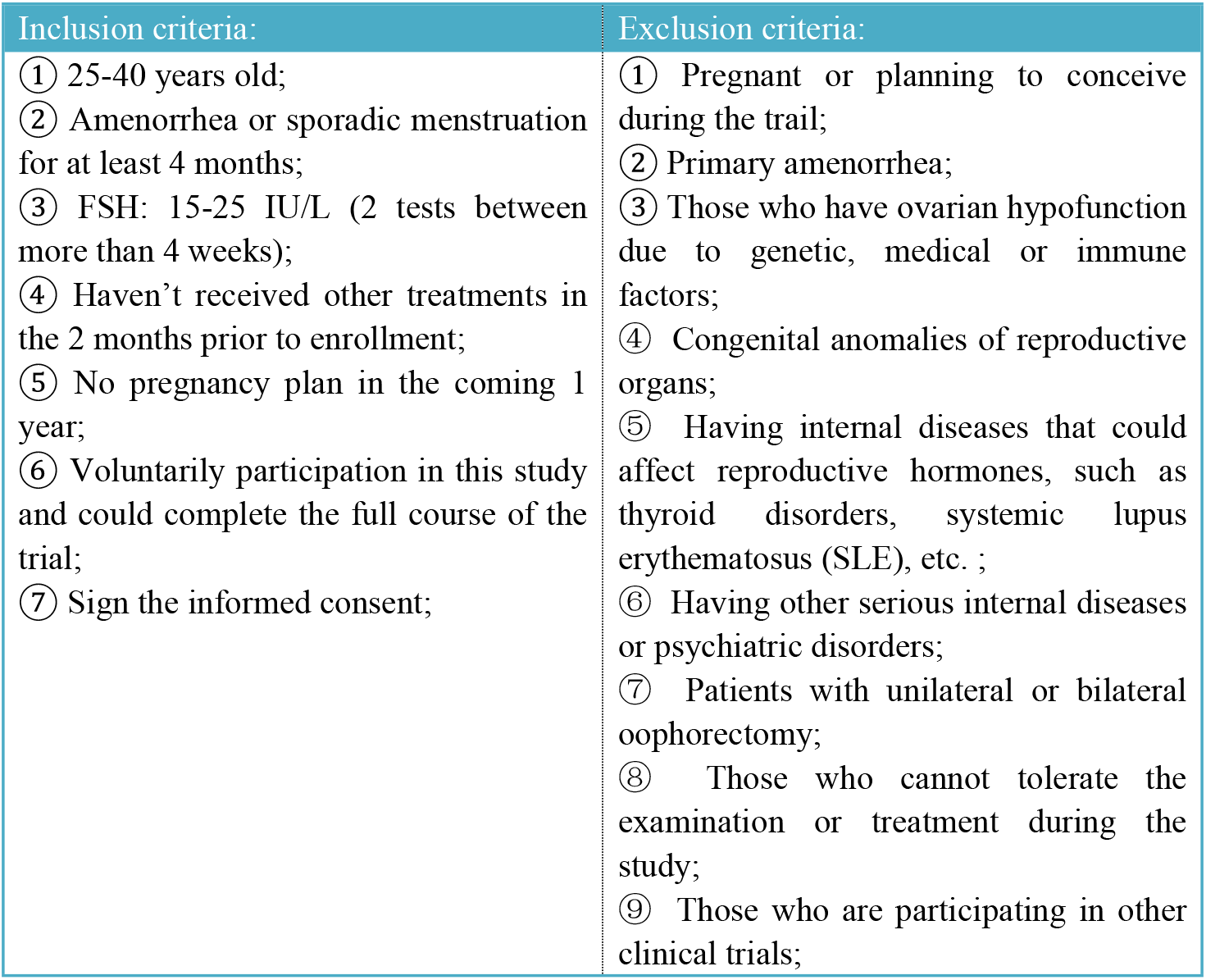
The inclusion criteria and exclusion criteria of subclinical POI patients

| Inclusion criteria: | Exclusion criteria: |
| --- | --- |
| ① 25-40 years old;<br>② Amenorrhea or sporadic menstruation for at least 4 months;<br>③ FSH: 15-25 IU/L (2 tests between more than 4 weeks);<br>④ Haven't received other treatments in the 2 months prior to enrollment;<br>⑤ No pregnancy plan in the coming 1 year;<br>⑥ Voluntarily participation in this study and could complete the full course of the trial;<br>⑦ Sign the informed consent; | ① Pregnant or planning to conceive during the trail;<br>② Primary amenorrhea;<br>③ Those who have ovarian hypofunction due to genetic, medical or immune factors;<br>④ Congenital anomalies of reproductive organs;<br>⑤ Having internal diseases that could affect reproductive hormones, such as thyroid disorders, systemic lupus erythematosus (SLE), etc. ;<br>⑥ Having other serious internal diseases or psychiatric disorders;<br>⑦ Patients with unilateral or bilateral oophorectomy;<br>⑧ Those who cannot tolerate the examination or treatment during the study;<br>⑨ Those who are participating in other clinical trials; |

### Randomization and allocation

Subjects will be randomly assigned to the EA group, the TEAS group or the waiting for treatment group with a 1:1:1 allocation. Each participant was assigned an identification number at the time of enrollment. The randomization sequence will be generated by SPSS 25 (Statistical Product and Service Solutions 25) statistical software and be randomly divided into three groups. Randomizers will contact participant via mobile text messages, informing them of the time and room of treatment and receiving participant confirmation. The grouping information will be stored in a computer with a password. Recruitors, randomizers, therapists, evaluators, and statisticians selected are different personnels.

### Interventions

The baseline lasts for two months, the treatment lasts for three months, and the follow-up lasts for twelve months. Participants in the former two groups will receive either EA or TEAS treatment for one hour (supine position for 30 minutes and prone position for 30 minutes). The treatment will be given three times a week during the first month, twice a week during the second month, and once a week during the third months. The treatment will be uninterrupted during menstruation. The treatment plan will be strictly carried out by two trained acupuncturists to ensure that the proper acupoints are stimulated and trial protocol is precisely followed. In addition to being given a paper-based schedule, all the patients will also be verbally reminded of the next treatment to ensure therapeutic effect. All participants will be discouraged from receiving any additional treatments for subclinical POI, including medications, traditional Chinese medicine, and other non-drug therapies. Subjects will be excluded if they receive less than 50% of the total treatments. Any treatment, symptom changes and relevant conditions administered by the patient during the trial should be filled out completely on the case report forms (CRFs). The acupuncture treatment is accordance with the Standards for Reporting Interventions in Controlled Trials of Acupuncture (STRICTA) checklist[25] **(Supplementary file 3)**.

#### EA group

The acupuncture protocol is based on the “regulating menstruation and promoting pregnancy” acupoints therapy which has been implemented by the Acupuncture and Moxibustion Hospital of China Academy of Chinese Medical Sciences (CACMS) [26]. We will adopt filiform stainless steel needles for one-time use (size: 0.25 mm × 40 mm and 0.30 mm × 75 mm, Huatuo brand, Suzhou Medical Appliance, Jiangsu Province, China) and electro-acupoint stimulators (SDZ-IIB, Huatuo brand, Suzhou Medical Appliance, Jiangsu Province, China) as treatment tools. The acupoints are: ① GV20 (Bai hui), GV24 (Shen ting), CV4 (Guan yuan) and bilateral GB13 (Ben shen), ST25 (Tian shu), KI12 (Da he), EX-CA1 (Zi gong), ST36 (Zu san li), SP6 (San yin jiao), KI3 (Tai xi), LR3 (Tai chong) and ② bilateral BL23 (Shen shu) and BL32 (Ci liao). The points in the two groups will be both used in each treatment. The positions and manipulation of the above acupoints[27] and listed in **Table 3** and **Figure 2**.

**Table 3.**
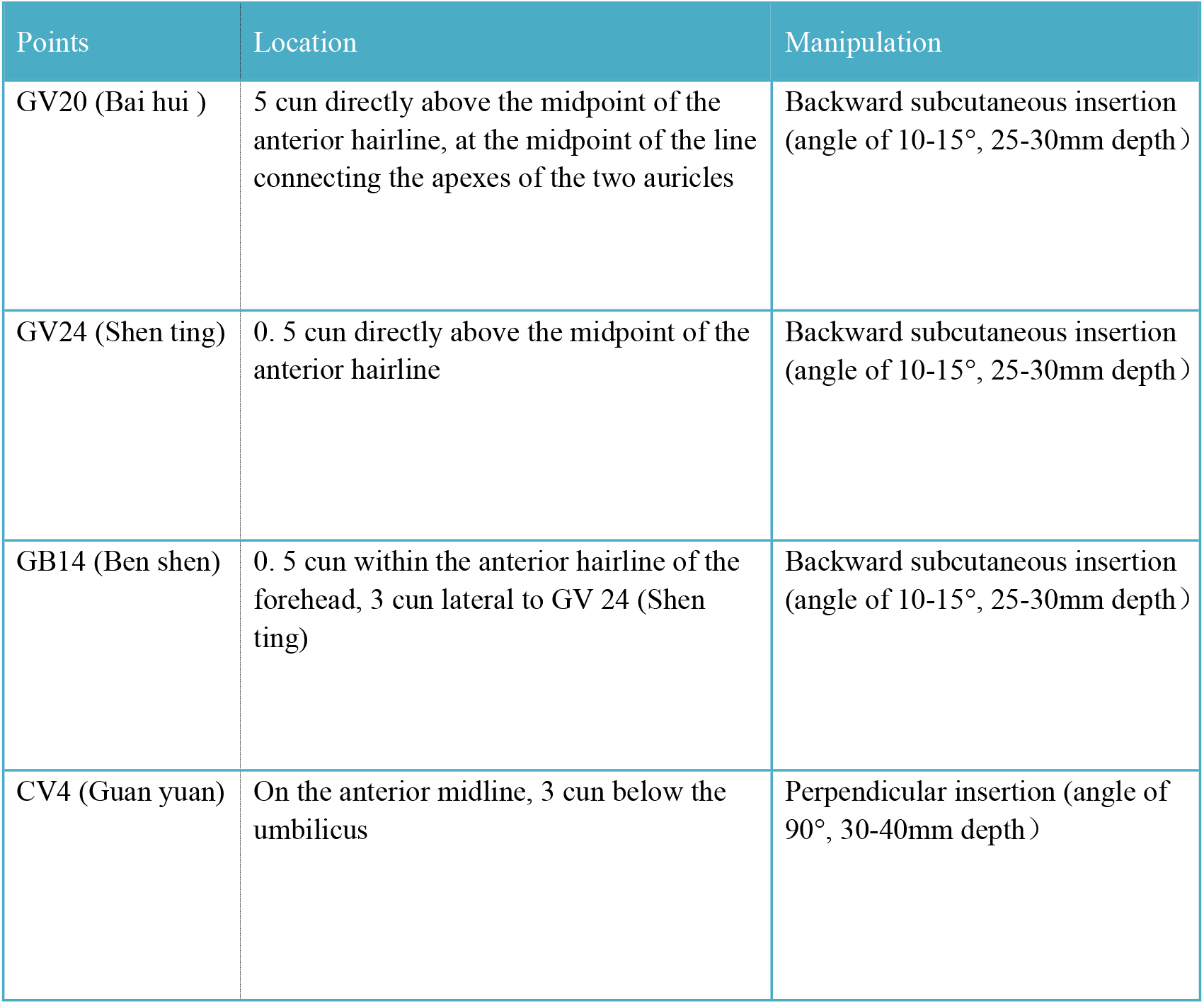
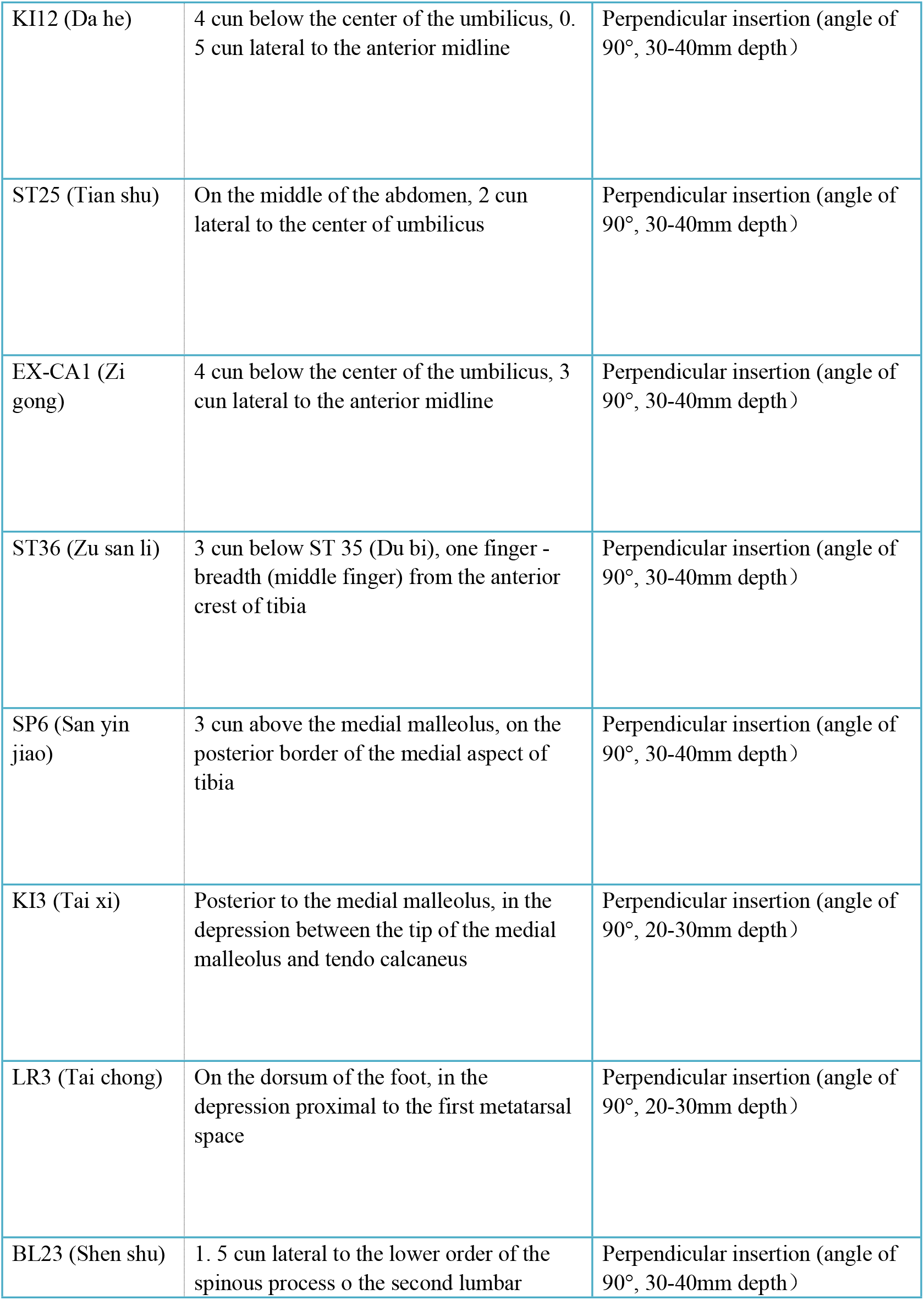
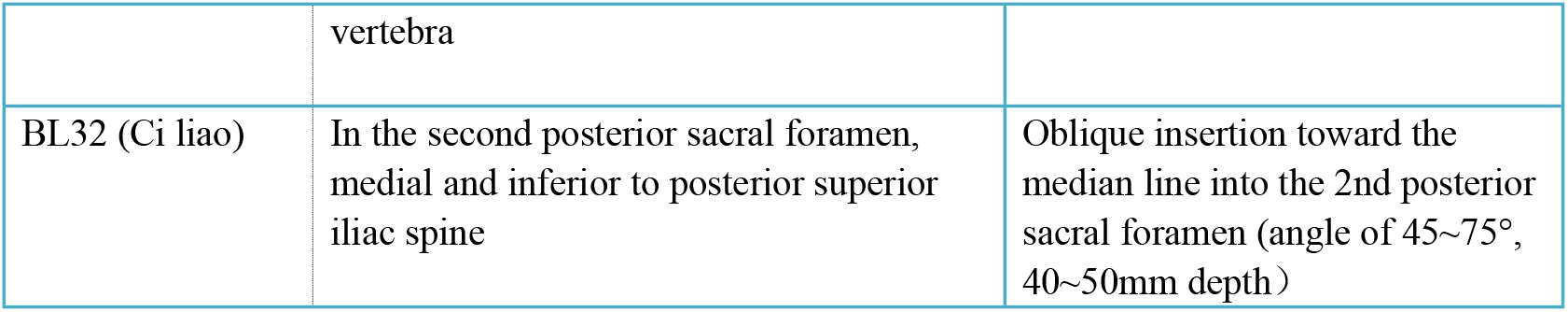
Locations and manipulation of the points

**Figure 2:**
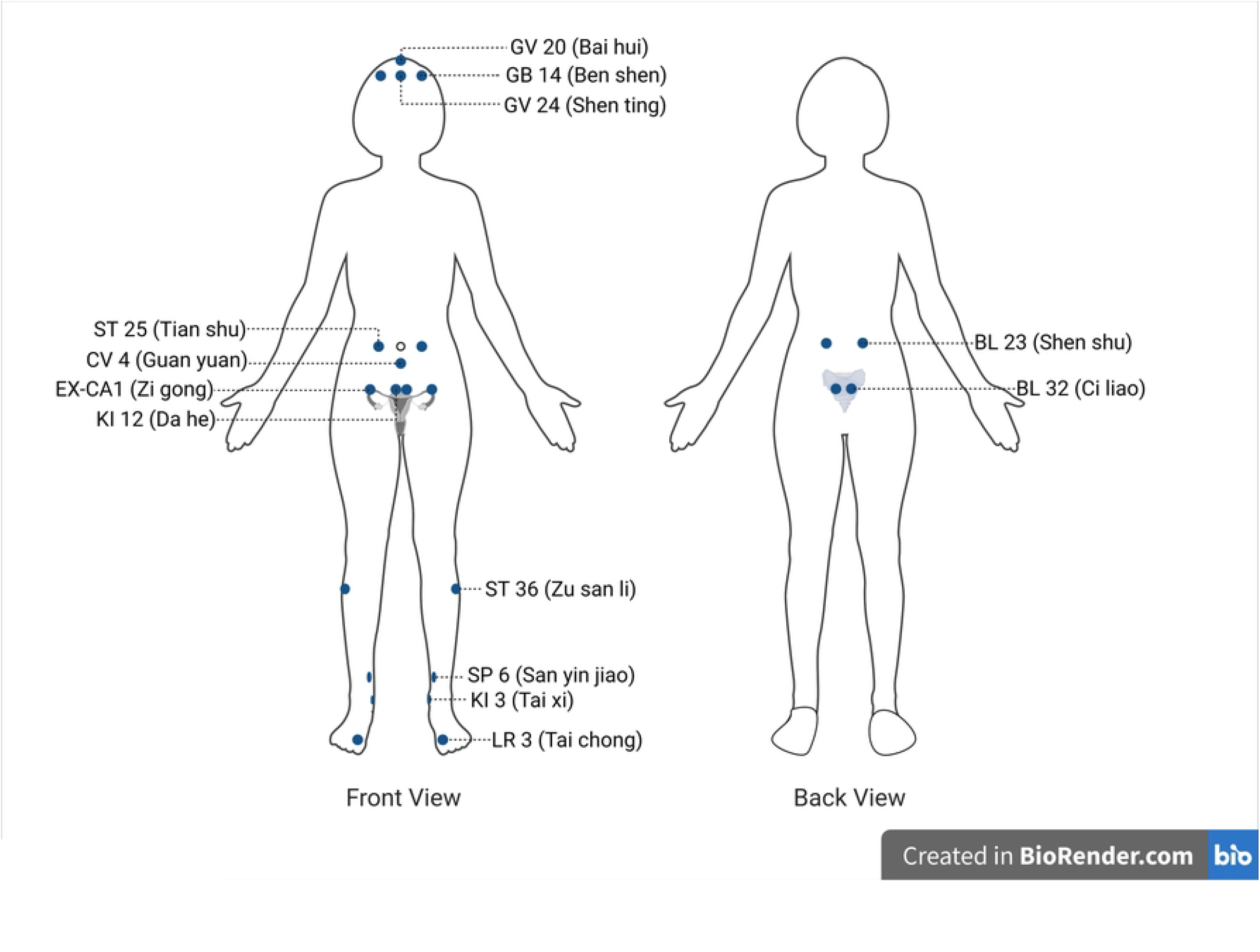

Acupuncture operations: In supine position, needles (0.25 mm × 40 mm) will be inserted in the following manner: Backward flat insertion 25 ∼ 30 mm in GV20 (Bai hui), GV24 (Shen ting) and GB13 (Ben shen). Straight insertion 30 ∼ 40 mm in ST25 (Tian shu), CV4 (Guan yuan), KI12 (Da he), EX-CA1 (Zi gong) and SP6 (San yin jiao). Straight insertion 20 ∼ 30 mm in KI3 (Tai xi) and LR3 (Tai chong). “De Qi” (“De Qi” is a feeling of sore, bloated, heavy or electric shock in needlepoint, which is the considered to be the basis for effectiveness) will be required on all acupoints. After obtaining “De Qi”, EA will be added with continuous waves on with frequency of 2Hz and intensity 3∼5 mA on bilateral ST25 (Tian shu) and EX-CA1 (Zi gong), to a level which can be tolerated by the patient. After 30 minutes, the patient will be asked to lie prone. The acupuncturist will insert BL23 (Shen shu) straight 30∼40 mm with acupuncture needles (0.25 mm × 40 mm), and then insert BL32 (Ci liao) obliquely toward the median line into the second posterior sacral foramen 40∼50 mm with acupuncture needles (0.30 mm × 75 mm). After obtaining “De Qi”, EA will be added on bilateral BL23 (Shen shu) and BL32 (Ci liao) with continuous waves on with frequency of 2Hz and intensity 3∼5 mA, to a level which can be tolerated by the patient. After 30 minutes, remove the EA and needles and the treatment comes to an end.

#### TEAS group

Patients in TEAS group will be treated with the same electro-acupoint stimulators in EA group (We will replace the clips at the end of the wire with factory-matched patches that can conduct electricity) on the following two groups of acupoints: ① bilateral EX-CA1 (Zi gong) and ST25 (Tian shu). ② bilateral BL23 (Shen shu) and BL32 (Ci liao). Points in the above two groups will be both used in each treatment.

TEAS operation process: Patients will first be asked to take supine pose. Electrode patches will be attached to the corresponding acupuncture points: a pair of positive and negative poles for bilateral ST25 (Tian shu) and EX-CA1 (Zi gong) respectively, and current will be set as 20∼30 mA in intensity and the same waveform, frequency as which in EA group. Then, the patient will be asked to lie prone. Bilateral BL23 (Shen shu) and BL32 (Ci liao) will be treated with same electricity mode. Acupoints in each group will be stimulated for 30 minutes.

#### Waiting for treatment group

No treatment will be given to the patients in waiting for treatment group. After the trial, patients in this group will be provided with the dominant treatment.

### Outcome measures

Primary outcomes will be the serum AMH and FSH. These levels will be measured by taking venous blood from the elbow on an empty stomach between 9 ∼ 11 am on day 2 ∼ 3 of the menstrual period for participants with regular menstruation. For patients with irregular menstruation or amenorrhea, the blood test will be taken between 9 ∼ 11 am on any day in the evaluation month.

Secondary outcomes will include: serum E_2_, LH, FSH/LH ratio, the menstrual status assessment, and the modified Kupperman Menopausal Index. The blood test indicators will be tested simultaneously with primary outcomes. These serological tests will be performed at our hospital by a pre-contacted examiner and a reliable testing machine. The menstrual status assessment refers to The International Federation of Gynecology and Obstetrics (IFGO) Recommendations on Terminologies and Definitions for Normal and Abnormal Uterine Bleeding[28], as defined by the following parameters. The modified Kupperman Menopausal Index is widely used internationally, and its role in clinical practice is well established[29-31]. Perimenopausal symptoms have different basic scores and degree scores, helping to rank their impact. The modified Kupperman Menopausal Index is shown in **Table 4**. Coordinators will inform subjects prior to the tests and help them get prepared for the tests.

**Table 4.** The Modified Kupperman Menopausal Index

| Symptoms | Basic score | Degree score |  |  |  | Symptom score |
| --- | --- | --- | --- | --- | --- | --- |
|  |  | 0 | 1 | 2 | 3 |  |
| Sweating and hot flashes | 4 | None | <3 times per day | 3-9 times per day | ≥10 times per day |  |
| Paresthesia | 2 | None | Weather-related | Frequent cold, hot, painful and numb feelings | Loss of hot, cold and pain sensation |  |
| Insomnia | 2 | None | Occasionally | Often and sleeping pills is effective | Having disruption to life |  |
| Nervousness | 2 | None | Occasionally | Often and self-controllable | Frequent and difficult to control |  |
| Melancholia | 1 | None | Occasionally | Often and self-controllable | Frequent and difficult to control |  |
| Vertigo | 1 | None | Occasionally | Often but no disruption to life | Having disruption to life |  |
| Fatigue | 1 | None | Occasionally | Climbing to the fourth floor is difficult | Restricted daily activities |  |
| Arthralgia myalgia | 1 | None | Occasionally | Often but no disruption to function | Having disruption to function |  |
| Headache | 1 | None | Occasionally | Often but tolerable | Need to be treated |  |
| Palpitation | 1 | None | Occasionally | Often but no disruption to life | Need to be treated |  |
| Formication | 1 | None | Occasionally | Often but tolerable | Need to be treated |  |
| Urinary tract infection | 2 | None | Occasionally | $\geq 3$ times a year and can recover itself | $\geq 3$ times a month and need to take medicine | |
| Sexual complaints | 2 | Normal | Decreased sexual desire | Painful intercourse | Loss of sexual desire |  |
| Total score | / |  |  |  |  |  |
| Evaluation of the degree | Normal Mild Moderate Severe |  |  |  |  |  |
|  | Symptom score = basic score * degree score. The total score is the sum of symptom scores.<br>The total score is less than 6 for normal, 6-15 for mild, 16-30 for moderate, and more than 30 for severe. |  |  |  |  |  |

### Safety

Adverse events (AEs), which include hemorrhage, dizziness, stagger needle, bent needle and unbearable pain will be dealt with in a timely manner by acupuncturists. In the meanwhile, the date, degree, treatment and duration of AEs will be recorded by the investigator on the CRFs. In case of serious AEs, the steering committee and ethics committee of hospital will be informed in the first instance and the acupuncturist will determine whether researcher team should exclude the participant from the study. We are responsible for the cost of any medication and care required in treatment of injury caused by this trial.

### Quality control and data management

The study protocol developers consulted extensively with gynecologists, acupuncturists, and statisticians in the each modification of the protocol. Prior to recruitment, coordinators underwent training to ensure they are well informed about the program. Recruiting sessions will be done by professional gynecologists and acupuncturists specializing in gynecology together. The data management committee, which is composed of experts from different departments and has no conflict of interest with the project, will monthly monitor the record of CRFs.

### Sample size calculation

This is an exploratory study without previous research for reference. Therefore, we adopted the State Food & Drug Administration’s (SFDA) minimum number of cases requirement of 30 as the number of participants in the trial group[32]. We intended to include 38 participants considering a loss rate of roughly 20%.

### Statistical analysis

Data will be double entered by two people in Microsoft Excel spreadsheet (Microsoft Corporation). Statistical analysis will be implemented by blinded bio-statisticians from an independent third party on SPSS25. Missing values will be handled using the mixed model for repeated technique. Continuous data will be expressed as mean and standard deviation. Repeated measurement data will be analyzed by repeated measurement data’s ANOVA test. First, Mauchly’s test of sphericity will be performed. If the data meets the H-F condition (spherical test *p* > 0.05), then ANOVA will be used. If the data does not meet the condition, the biostatisticians will calculate the spherical symmetric coefficient corrected’s degree of freedom by the unary F statistic to obtain the probability of correction. The Chi-square test will be used on categorical data. The rank-sum test will be used on ranked data. A two-sided p < 0. 05 will be considered statistically significant for all analyses.

## DISCUSSION

POI, one of the main causes of female infertility, brings great physical and mental suffering to patients and has recently won increasing attention. Seventy to ninety percent of POI is caused by unspecified etiology. Given the fact that POI is a slowly developing disease, finding an appropriate method to slow down the progression of the disease in patients has merit. Although HRT is widely performed in the treatment of POI, its effect is limited by unavoidable side effects. Acupuncture, a long-established and accessible therapy, has become a potential treatment for halting the decline of ovarian function in women of childbearing age. The reason why we designed this trial is to investigate the efficacy and safety of acupuncture (including EA and TEAS) for treating subclinical POI.

### The Strengths of the Study

First, acupuncture relevant treatments are promising methods for the treatment of subclinical stage POI patients. On the one hand, because of the drug abuse and surgical damage, a large number of people tend to advocate green therapies including acupuncture, exercise, and lifestyle changes as their first choice for treatment. Although these therapies are less effective than drugs and surgery in many organic diseases, they have unique advantages in treating functional, chronic, physical and mental disorders, mostly with a holistic treatment effect and high therapeutic efficiency. On the other hand, moderate stress can initiate endogenous protective mechanisms in the body, producing broad-spectrum, non-specific regulatory effects to prevent the onset and progression of disease. To some extent, acupuncture can be considered as a stressor that generates benign stress. Related studies have shown that acupuncture can regulate the release of stress hormones such as corticotropin-releasing hormone (CRH), β -endorphin (β -EP), and adrenocorticotropic hormone (ACTH)[33-34]. Therefore, long-term preventive treatment on POI with acupuncture is becoming clinically possible.

Second, the treatment plan we selected is not only well supported by traditional Chinese medical theory **(Supplementary file 4)**, but also has been proven to have an ideal effect on POI and relevant gynecological diseases including: diminished ovarian reserve, premature ovarian failure and pregnancy assistance[14,26,35-36]. According to modern research, the mechanisms of acupuncture for POI mainly lies in regulating reproductive endocrine hormones, improving ovarian morphology and blood flow, improving neurological function, and relieving mental stress. Zhang and Wang[37-38] revealed that EA can regulate hormone levels in premature ovarian failure rats by up-regulate PI3K/Akt signaling pathway, and reduce the apoptosis of granulosa cells. Stener-Victorin[39] found that improving blood flow to the ovaries is one of the mechanisms by which acupuncture treats gynecological diseases. Zhou[19] conducted that reproductive hormone modulation effects may be caused by the effects of acupuncture in the autonomic system.

### Limitations

First, the sample size of the clinical trial is small. However, this trial has met the minimum sample size for clinical trials of SFDA and is sufficient to draw preliminary conclusions. In order to obtain a reliable result, we will attempt to reduce the dropout rate during the process of the trial through excellent communication, clinic follow-up reminders, accessibility to patient concerns and questions, and reasonable patient accommodation where applicable.

Additionally, we didn’t choose sham acupuncture as our control group because a large number of people have experienced acupuncture treatment in China, making it difficult to deceive patients with sham acupuncture. In place of sham acupuncture, we selected waiting for treatment group as our control measurement because it is consistent with clinical practice.

## Conclusion

Since there are no ideal treatment plan for subclinical POI. We expect this randomized controlled trial demonstrate whether EA or TAES will be the ideal treatment for subclinical POI on regulating hormone levels, improving clinical symptoms and delaying disease progression. Another obvious deficiency of this study is that we can only prove whether the this EA treatment plan and TEAS treatment plan selected in the study have significant clinical efficacy, but can not clarify wheter EA or TEAS is better for subclinical POI treatment for the number of points selected is different. We may keep working on this issue in the subsequent research.

## Data Availability

No datasets were generated or analysed during the current study. All relevant data from this study will be made available upon study completion.

http://www.chictr.org.cn/showproj.aspx?proj=125026.

## DECLARATION

### Ethics approval and consent to participate

This study will be conducted in compliance with the norms governing research involving human subjects, following approval from the Ethics committees of the Third Affiliated Hospital of Zhejiang Chinese Medical University under Ethics approval numbers: ZSLL-KY-2021-004-01. The participants will be given clarifications concerning the objectives, procedures of the study and will be asked to sign a statement of informed consent prior to the study beginning, both in writing and orally.

### Consent for publication

The results will be published in peer-reviewed and open-access journals.

### Availability of data and materials

All data will be available upon reasonable request.

### Competing interests

The authors declare that they have no competing interests.

### Funding

This trial was funded by the Administration of Traditional Chinese Medicine of Zhejiang Province (NO. 2021ZB145) and the Inheritance Studio Construction Project of National Famous Old Chinese Medicine Experts of Dr. Wang Zhanglian (Letter of the Department of Personnel and Education of the National Administration of Traditional Chinese Medicine, No.(2022)75). **(Supplementary file 5)**

### Authors’ contributions

LFC is responsible for this study and designed the trial protocol. JLW and LLD drafted the manuscript. All of the other authors made substantial contributions to the design of this study or revising the manuscript critically.

### Trial status

The trial is currently enrolling participants. Enrollment and completion of the trial are scheduled from May 1^st^, 2021 to August 31^th^, 2024.

## Abbreviations

POI: Premature ovarian insufficiency
EA: electro-acupuncture
TEAS: transcutaneous electrical acupoint stimulation
HRT: hormone replacement therapy
FSH: follicle-stimulating hormone
AMH: anti-Müllerian hormone
E_2_: estradiol
LH: luteinizing hormone
CRF: case report form
AEs: adverse events

## Notes

### Competing Interest Statement

The authors have declared no competing interest.

### Clinical Trial

Trial was registered in the Chinese Clinical Trail Registry (ChiCTR-2100045598).

### Funding Statement

Dr. Lifang Chen and Dr. Zhanglian Wang are initials of the authors who received the awards. Dr. Lifang Chen received fund from the Administration of Traditional Chinese Medicine of Zhejiang Province (NO. 2021ZB145) in 2020. Dr. Zhanglian Wang received fund from the Inheritance Studio Construction Project of National Famous Old Chinese Medicine Experts of Dr. Wang Zhanglian (Letter of the Department of Personnel and Education of the National Administration of Traditional Chinese Medicine, No.(2022)75) in 2022. The URL of the former funder is http://www.zjtcm.gov.cn/. The URL of the latter funder is http://www.satcm.gov.cn/. The funders had and will not have a role in study design, data collection and analysis, decision to publish, or preparation of the manuscript.

### Author Declarations

the Ethics committees of the Third Affiliated Hospital of Zhejiang Chinese Medical University.Ethics approval numbers: ZSLL-KY-2021-004-01

